# Transcutaneous auricular vagus nerve stimulation demonstrates effects on pupil responses in Parkinson’s disease

**DOI:** 10.64898/2026.09.13.26362895

**Authors:** L. M. Schmidt, J. Stalter, C. M. Thiel, K. Janitzky, K. Witt

**Affiliations:** Department of Neurology, School of Medicine and Health Sciences, Carl von Ossietzky University, Oldenburg, Germany; University Clinic for Neurology at the Evangelical Hospital, Oldenburg, Germany; Biological Psychology Lab, Department of Psychology, School of Medicine and Health Sciences, Carl von Ossietzky University, Oldenburg, Germany; Research Center Neurosensory Science, Carl von Ossietzky University Oldenburg, Oldenburg, Germany; Cluster of Excellence "Hearing4all", Carl von Ossietzky University Oldenburg, Oldenburg, Germany

**Keywords:** taVNS, pupillometry, Parkinson’s disease, LC-NE system

## Abstract

**Background:** The locus coeruleus-norepinephrine (LC-NE) system is affected early in Parkinson’s disease (PD) and is associated with various non-motor symptoms. Transcutaneous auricular vagus nerve stimulation (taVNS) is a promising non-invasive approach for improving PD symptoms, and it has been hypothesized that its action is linked to the LC-NE brainstem system; however, its neurophysiological mechanisms remain poorly understood.

**Objective:** Given the close relationship between LC-NE activity and pupil dynamics, pupillometry may provide a non-invasive measure of LC-related activity during taVNS. This monocentric, single-blinded, sham-controlled study investigated the impact of taVNS on pupillary response as an indicator of taVNS effects at the level of the brainstem.

**Methods:** Twenty-one patients with PD and 18 age-matched healthy controls were included. Real and sham stimulation were applied in a randomized cross-over design, and pupil size was continuously recorded. A local-global auditory processing task, which elicits arousal-related responses to auditory irregularities, was incorporated as a control condition to evaluate pupil responsiveness associated with LC-NE system activity.

**Results:** Real taVNS elicited greater pupil dilation than sham stimulation, with a significant effect emerging 2710–3640 ms after stimulation onset in patients with PD. In contrast, no significant difference was found between real and sham stimulation in healthy controls. The local-global auditory processing task elicited larger pupil responses to deviant than non-deviant stimuli in both groups, indicating intact LC-NE reactivity.

**Conclusion:** TaVNS elicited a significant pupillary response in individuals with PD but not in healthy older adults, providing evidence of physiological responsiveness to taVNS in PD.

## 1 Introduction

In PD, degeneration of the locus coeruleus-norepinephrine (LC-NE) system has been linked to a range of non-motor symptoms, such as sleep disorders, neuropsychiatric symptoms, and autonomic and cognitive dysfunction (Paredes-Rodriguez et al., 2020; Solopchuk et al., 2018). The LC is a brainstem nucleus with widespread projections throughout the brain and plays an important role in arousal, attention, and cognition through NE release (Benarroch, 2009, 2018; Chandler et al., 2014). Importantly, LC pathology is thought to occur in early phases of PD, potentially even preceding the degeneration of dopaminergic neurons mainly associated with well-known motor symptoms of the disease (Braak et al., 2004). Therefore, the LC-NE system has increasingly been considered a potential target for therapeutic interventions to improve non-motor symptoms in PD (Proietti et al., 2025).

TaVNS has emerged as a potential non-invasive neuromodulatory approach for PD. By stimulating the auricular branch of the vagus nerve, taVNS engages the nucleus tractus solitarius, the primary brainstem relay for vagal afferent input (Frangos et al., 2015; Kraus et al., 2013; Yakunina et al., 2017). From this nucleus, signals are transmitted to several brain regions, including the LC, thereby modulating the LC-NE system and its projection areas (Dietrich et al., 2008; Frangos et al., 2015). Recently, more studies on the effect of taVNS in PD have been published (see Eissazade et al., 2025; Golzarian et al., 2025; Proietti et al., 2025 for detailed reviews). Sham-controlled studies have reported improvements in selected gait parameters and some non-motor outcomes, including reaction time and anxiety, whereas effects on broader clinical measures such as the MDS-UPDRS-III have been absent or inconsistent (Marano et al., 2022, 2024; Van Midden et al., 2024; Zhang et al., 2023). Despite promising clinical findings, the neurophysiological mechanisms underlying the effects of taVNS in PD remain poorly understood. In particular, establishing whether taVNS engages the LC-NE system in PD and how such engagement may be reflected in physiological measures is an important step towards understanding its mechanism of action and therapeutic potential.

LC neurons operate in tonic and phasic modes, reflecting ongoing arousal and brief responses to salient or task-relevant events, respectively, thereby contributing to the regulation of attention and behaviour (Aston-Jones & Cohen, 2005; Rajkowski et al., 1994). Pupil size is closely coupled to LC activity and varies with arousal and cognitive processes (Joshi & Gold, 2020). Baseline pupil size has been associated with tonic LC activity and arousal, whereas transient pupil dilations are commonly interpreted as phasic responses to salient or task-relevant events (Joshi et al., 2016; Joshi & Gold, 2020). In monkeys, LC activity precedes changes in pupil size, both during spontaneous fluctuations in pupil size and in response to unexpected auditory events (Joshi et al., 2016). Similarly, auditory deviants elicited responses in both the LC and pupil size in humans (Mazancieux et al., 2023). Pupil dynamics provide a non-invasive marker of neuromodulatory or arousal-related activity that is closely associated with the LC-NE system but also reflects other influences, including cholinergic activity, autonomic state, arousal and sensory processing (Gilzenrat et al., 2010; Liu et al., 2017; Murphy et al., 2014; Reimer et al., 2016). A taVNS-induced pupil response may provide indirect evidence compatible with recruitment of ascending neuromodulatory systems, including LC-NE pathways (Farmer et al., 2021; Ludwig et al., 2021).

Consistent with this interpretation, previous studies have linked vagal stimulation to changes in pupil size and LC activity. In rats, invasive VNS has been associated with pupil dilation and LC activation in parallel (Bianca & Komisaruk, 2007; Collins et al., 2021; Mridha et al., 2021). In young healthy individuals, several studies have reported increased pupil dilation during taVNS (Lloyd et al., 2023; Sharon et al., 2021; Urbin et al., 2021; Wienke et al., 2023), whereas others have found no significant effect of taVNS on pupil size (Burger et al., 2020; D’Agostini et al., 2021, 2022; Keute et al., 2019). Importantly, a recent meta-analysis found that the pupillary response differed according to stimulation protocol, with pulsed taVNS (i.e., 0.5 - 3 s) showing evidence for increased pupil dilation, whereas conventional taVNS (e.g., 30 s of stimulation) showed evidence for a null effect (Pervaz et al., 2025).

Whether these pupillary responses translate to PD, in which the LC is affected by disease-related changes, remains unknown. In particular, it is unclear whether LC-NE system alterations in PD influence pupillary response to taVNS. Demonstrating a taVNS-induced pupillary response in PD would provide a first step towards establishing whether taVNS engages LC-NE-related processes in PD and could help understand its potential therapeutic effects. The present study therefore investigated whether taVNS elicits a pupillary response in individuals with PD and healthy older adults. We hypothesized that real taVNS would lead to greater pupil dilation than sham stimulation and that the magnitude of taVNS-induced pupil response would differ between groups. A local-global auditory processing task was also incorporated in this study, which is recognized for eliciting arousal-related responses to breaches of temporal auditory regularities. This task is known to activate the LC-NE system and was utilized as a control condition to evaluate pupil responsiveness associated with LC-NE system activation (Mazancieux et al., 2023).

## 2 Methods

This prospective, monocentric, single-blinded and sham-controlled study was conducted at the University Department of Neurology outpatient clinic at the Evangelisches Krankenhaus Oldenburg, Germany. The study protocol was reviewed and approved by the Medical Ethics Committee of Carl von Ossietzky University Oldenburg (file number 2025-121) and preregistered in the German Register for Clinical Studies (DRKS00037865). The data were collected between October 2025 and July 2026.

### 2.1 Participants

Participants were recruited for the PD group if they had a clinical diagnosis of idiopathic Parkinson’s disease according to the Movement Disorder Society Clinical Diagnostic Criteria for PD (Postuma et al., 2015). Patients were recruited through the Dept. of Neurology of the Evangelic Hospital Oldenburg, while healthy controls were recruited through flyers and a website. Exclusion criteria for all participants included any neurological or psychiatric disease (except PD), diabetes mellitus, or relevant ocular conditions (i.e., previous eye surgery, glaucoma, pseudoexfoliation syndrome, corneal disease, nonspherical pupils, or clinically relevant anisocoria). Further exclusion criteria were major cognitive decline (participants scored less than 21 points on the Montral Cognitive Assessment, MoCA, Nasreddine et al. 2014), the use of centrally acting medications (except PD medication), based on previously described criteria (Kelbsch et al., 2019), as well as the use of a deep brain stimulation device or a visual aid incompatible with pupil tracking (e.g., multifocal contact lenses, or reflective glasses). Participants using removable visual aids were included if their visual acuity was sufficient to maintain fixation on a cross during pupil data collection.

Given the absence of data on taVNS-induced pupillary reactivity in older adults or patients with PD, the effect size assumption was derived from a meta-analysis of pupillary reactivity following taVNS in healthy individuals (Pervaz et al., 2025). While the pooled effect size across studies using pulsed taVNS was small-to-moderate (*d* = 0.356, 95% CI [0.186, 0.528]), individual studies showed considerable heterogeneity (*τ* = 0.089, 95% CI [0.000, 0.272]). Given the variability in reported effect sizes and the proof-of-concept nature of our design, we pragmatically adopted a comparatively optimistic between-group effect size of *d* = 1.1, reflecting the upper range of effect sizes reported across individual studies in the meta-analysis (Pervaz et al., 2025). Sensitivity analysis was performed using a two-sided Mann-Whitney U test across a range of assumed effect sizes. For *N* = 30 (minimum 15 participants per group), the analysis indicated 80% power at α = 0.05 for the assumed between-group effect size. To mitigate potential data loss due to artifacts or technical issues, additional participants may be recruited, with 15 participants per group representing the lower threshold.

### 2.2 Procedure

Each participant underwent both stimulation conditions (real and sham), with the order of conditions randomly assigned. The participants were blinded to the electrode position expected to stimulate the auricular branch of the vagus nerve. The study was conducted in a single session, which was scheduled in the morning whenever possible. Participants with PD were assessed during regular dopamine replacement therapy (med-on). All participants provided written informed consent prior to data collection. Participants received a compensation of 12€ per hour for their participation in the study.

Before the experiment, participants were asked to get sufficient sleep the night before and to refrain from consuming any nicotine or caffeine-containing products 4 hours prior to the experiment. Participants completed the MoCA and visual acuity was assessed using a visual-acuity chart. The Participants then completed a short questionnaire on their current status (e.g., hours of sleep the previous night). Additional questionnaires assessing demographics and baseline status, symptoms of depression (Beck’s Depression Inventory, BDI-II; Beck et al., 2011), health-related quality of life (Short Form 12, SF-12; Ware et al., 1996), and REM sleep behaviour disorder severity (International REM Sleep Behavior Disorder Symptoms Severity Scale, IR-REM-BD; Fantini et al., 2024) were completed. Participants subsequently completed two computer-based attention tasks (Divided Attention and Go/No-Go task, TAP-M; Zimmermann & Fimm, 2020).

Pupillometry was recorded continuously throughout the experiment, which consisted of two taVNS conditions (real, sham) and a separate auditory oddball task, which was included as a control condition to assess LC-NE-related pupil responses to salient auditory events. An overview of the experimental procedure shown in Figure 1.

**Figure 1.**
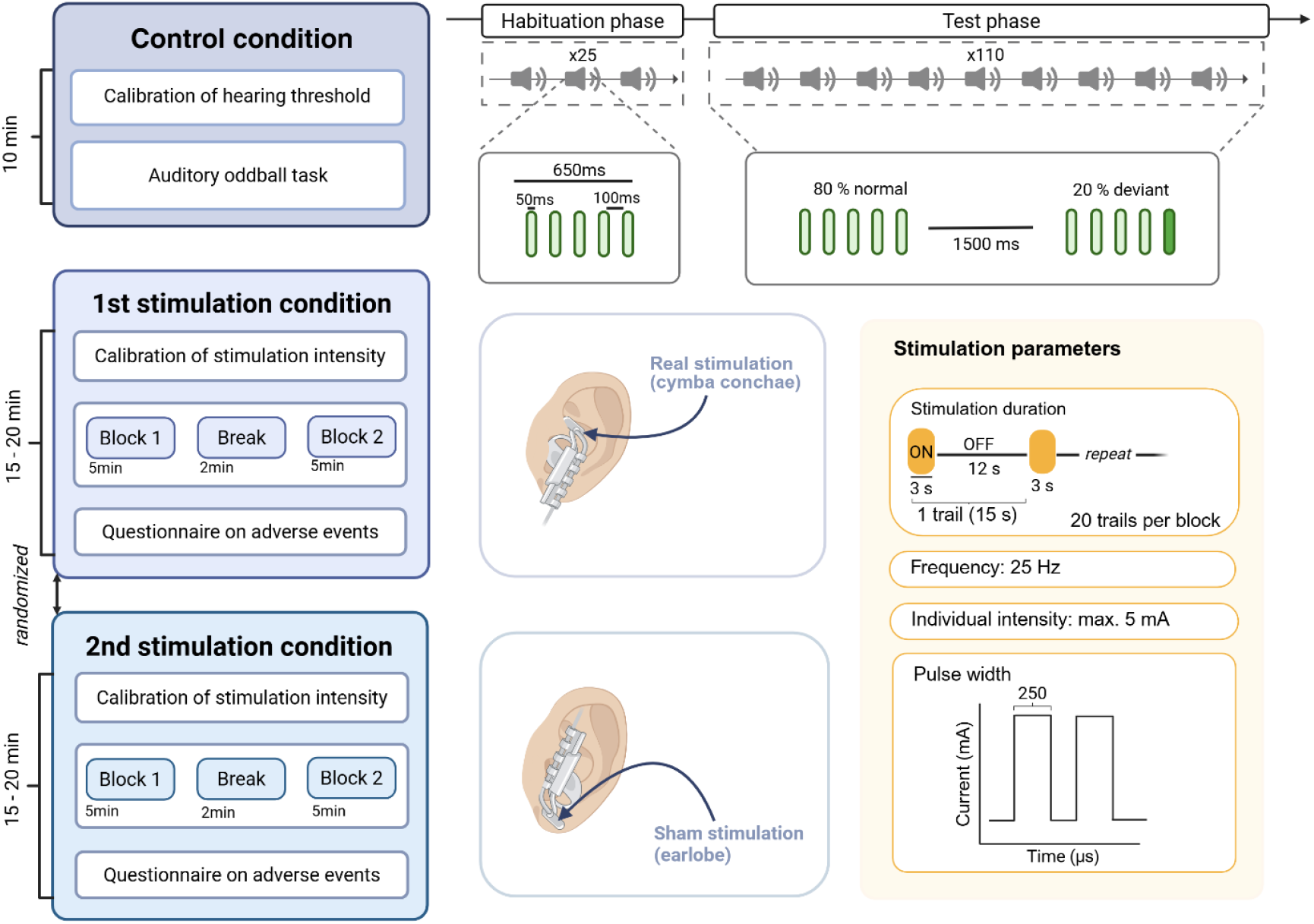
Overview of the experimental procedure. Participants completed the auditory oddball experiment (control condition), followed by two taVNS conditions. The figure also displays details of the local-global auditory processing task and stimulation parameters. Created in BioRender.com

Following the pupillometry experiment, patients with PD underwent disease-specific clinical assessments, including the Movement Disorders Society Unified Parkinson’s Disease Rating Scale (MDS-UPDRS; Goetz et al., 2008) and the Hoehn and Yahr (H&Y; Hoehn & Yahr, 1967) scale.

### 2.3 taVNS stimulation

#### 2.3.1 taVNS stimulation setup

The stimulation was controlled using Matlab (Matlab R2020b, Mathworks, Inc., Natick, MA, USA), which sent the digital stimulation signals through a digital-to-analog converter (USB-6341 DAQ; National Instruments, Austin, TX, USA) to a bipolar constant current stimulator (DS5 stimulator, Welwyn Garden City, UK). To connect the DS5 to the ear electrode, a custom-made adapter (converting 3.5 mm jack to double USB) was used. An ear electrode (legacy electrode, NEMOS^®^, Cerbomed, Erlangen, Germany), consisting of two titanium electrodes attached to a gel frame, was used. Before electrode placement, participants’ skin was cleaned with alcohol wipes and electrode pads were coated with electrode contact spray and attached to the electrodes to ensure better conductivity and stable contact to the skin. The electrode was held in position with an earpiece and further secured with medical tape.

#### 2.3.2 taVNS stimulation parameters

For both real (cymba conchae) and sham (earlobe) stimulation, the ear electrode was applied to the left ear. The left ear was selected in accordance with the conventional approach to vagus nerve stimulation, as left-sided stimulation is generally preferred due to concerns about potential cardiac effects associated with right-sided vagus nerve stimulation. Stimulation consisted of 3-second trains of rectangular pulses, with an inter-trial-interval of 12 seconds. Brief pulsed stimulation was selected because previous meta-analytic evidence suggests that short stimulation trains are more consistently associated with time-locked pupil dilation than conventional longer stimulation blocks (Pervaz et al., 2025). The other stimulation parameters included a pulse width of 250 μs, frequency of 25 Hz and a maximum stimulation intensity of 5 mA. More details on the stimulation parameters are displayed in Figure 1. Stimulation intensity was individually adjusted using a subjective sensory intensity scale ranging from 0 (no sensation) to 100 (painful sensation), with 80 representing a strong but non-painful sensation. During the calibration procedure, 5-second stimulation trains were delivered and participants rated each stimulation on the scale. Intensity was increased from 0.4 mA in 0.2-mA steps until a rating of 70 was reached, followed by 0.1-mA steps until just below 90. The intensity was then decreased until a rating of 70 was reached. This staircase procedure was repeated once, yielding four intensity values corresponding to a rating of 80. The mean of these values was used as the individual stimulation intensity.

#### 2.3.3 Adverse events of stimulation

The Adverse Events Questionnaire for taVNS (taVNS-AEQ) is a semi-structured questionnaire designed to assess the duration and severity of symptoms experienced during or after taVNS (Meier-Bartelt et al., 2026). It also assesses participants’ willingness to take part in future studies using the same stimulation protocol and their perception of whether they received real or sham stimulation.

### 2.4 Pupillometry

Pupil size was recorded monocularly from the left eye at 500 Hz using a desktop-mounted Eyelink 1000 (SR Research Ltd., 2005) running version 4.594 of the host software. Participants were seated in a comfortable, height-adjustable chair in front of the eyetracker and screen, with their chin and forehead resting on the Eyelink headrest (SR Research Head Support). The eyetracker was adjusted to ensure reliable pupil recording and was calibrated prior to data collection. The room was dimly lit during data recording. Pupil size was recorded throughout both experimental tasks.

#### 2.4.1 taVNS experiment procedure

The electrode was applied according to the randomly assigned stimulation condition (real or sham), and individual stimulation intensity was determined as previously described. Each stimulation condition lasted approximately 10 min and included a short break after the first 5 min to ensure participant comfort. During stimulation and simultaneous pupil recording, participants were instructed to fixate on a cross presented on the screen. After completion of the first stimulation condition, the electrode was removed and participants completed the taVNS-AEQ. Ten minutes after the end of the first stimulation condition, the second stimulation condition was administered following the same procedure.

#### 2.4.2 Local-global auditory processing task

As a control condition, a local-global auditory processing task was used to assess pupil dilation responses to deviant auditory stimuli. Stimulus presentation was controlled by Experiment Builder (SR Research Ltd., 2024). Before the task, the sound volume was individually adjusted to ensure that the tones were clearly audible and comfortable for each participant. The task was adapted from Mazancieux et al. (2023) and consisted of two different tones: a lower-pitched standard (non-deviant, “X”) tone and a higher-pitched deviant tone (“Y”). Specifically, each sound consisted of three sinusoidal components, the non-deviant stimulus comprised 350, 700, and 1400 Hz components, whereas the deviant stimulus comprised 500, 1000, and 2000 Hz components. Each trial consisted of a sequence of 5 tones. Of the 110 experimental trials, 80% consisted exclusively of standard tones (X-X-X-X-X), whereas in 20% of trials, the fifth tone was a deviant tone (X-X-X-X-Y). The complete trial duration was approximately 3.5 seconds, including a post-sequence inter-trial interval. Before the experimental trials, participants completed 25 habituation trials consisting exclusively of standard tones. The task lasted approximately 8.5 min. Participants were instructed to focus on a fixation cross on the screen and count the number of deviant tones to ensure attention to the auditory task.

#### 2.4.3 Pupil data analysis

Pupil data were pre-processed following the recommendations by Mathôt & Vilotijević (2022). Raw eye-tracking data were converted from .edf to .asc format and subsequently transformed into BIDS-compatible .tsv files. Data preprocessing and analysis were performed in RStudio version 4.4.3 (RStudio Team, 2020). To ensure blinding, each participant’s dataset was pre-processed without knowledge of the assigned stimulation condition.

The preprocessing pipeline consisted of the following steps. First, blinks were identified using a per-recording percentile threshold and removed with a ± 50 ms margin. Second, samples with an absolute pupil velocity above the 99.5^th^ percentiles (computed on a 3-point smoothed signal) were flagged as outliers and removed with a ± 20 ms margin. Pupil values, defined as falling below the 1^st^ and 99^th^ percentile of the blink-cleaned signal, were additionally removed. Third, missing data segments of ≤ 500 ms were linearly interpolated. Finally, the data were low-pass filtered using a 4th-order Butterworth filter with a cutoff frequency of 4 Hz. All datasets were visually inspected before and after preprocessing.

After preprocessing, data were segmented into epochs time-locked to stimulus onset. For taVNS, epochs ranged from -500 to 6000 ms relative to stimulation onset. For the local-global auditory processing task, epochs were centred on the fifth tone and ranged from -500 to 2500 ms. Trials were excluded if the baseline-window mean deviated more than 2 standard deviations (*SD)* from the participant’s own mean baseline (|*z*| > 2). Additionally, trials with substantial data loss (more than 50% of missing samples, more than 50% interpolated samples, or fewer than 50 valid samples in total) were excluded. Participants retaining fewer than 40% of the maximum trials were excluded from further analysis. Finally, pupil data were baseline corrected using the -500 to 0 ms pre-stimulus interval, computed separately for each trial.

### 2.5 Statistical analysis

All statistical analyses were conducted in RStudio (RStudio Team, 2020) version 4.4.3. Group differences in demographic and baseline characteristics were assessed using t-tests or Wilcoxon rank-sum tests. Differences in stimulation intensity (mA) between real and sham stimulation conditions within each group were assessed using paired-samples t-tests. Statistical significance was defined as *p* < 0.05 (two-sided).

To assess differences in pupil responses between experimental conditions (real vs. sham; deviant vs. non-deviant), cluster-based permutation analyses (Maris & Oostenveld, 2007) were conducted separately for healthy and PD groups. This approach was chosen over analysing a single summary value (e.g., peak pupil dilation) because it allows differences to be detected across the entire time course without an a priori assumption about when an effect occurs. Prior to analysis, epoched pupil data were resampled from 500 to 100 Hz, resulting in 10 ms time bins. For the taVNS experiment, difference scores were calculated for each participant and time bin; for the local-global auditory processing task, difference scores were calculated analogously. This enabled the analysis of the differences within each group for both the taVNS experiment (ΔPD_or ΔHC = real – sham) and the local-global auditory processing task (ΔPD or ΔHC = deviant – non-deviant).

At each time bin, a one-sample t-statistic (sign-flip approach) was calculated to test whether the difference scores differed from zero. Temporally adjacent time bins exceeding the critical t-value corresponding to a two-sided *α* = .05 (*t_crit* = 2.13, *df* = 15, for healthy controls; *t_crit* = 2.10, *df* = 18, for the PD group) were grouped into clusters. Cluster level-statistics were calculated as the sum of the absolute t-values within each cluster. These were compared against a null distribution generated from 10,000 sign-flipping permutations, thereby controlling the family-wise error rate across all time bins. To test whether the difference in taVNS or auditory processing differed between the healthy control and PD groups, two-sample cluster-based permutation tests were performed on the respective differences (ΔGroup = ΔHC – ΔPD). Welch’s t-statistics were calculated at each time bin, and temporally adjacent time bins exceeding the critical t-value corresponding to a one-sided α = .05 (*t_crit* = 1.68–1.72, Welch– Satterthwaite *df* = 22.2–33.0, varying across time bins) were grouped into clusters. Clusters were formed and evaluated as described above. Because the analyses compared two independent groups, a null distribution was generated by randomly shuffling group labels across 10,000 permutations. For all analyses, permutation-based p-values (+ 1 correction; (Phipson & Smyth, 2010) were used to determine cluster significance. Additionally, exploratory cluster-based permutation analyses were conducted separately for the healthy and PD groups to assess potential condition order effects by comparing the stimulation effects between participants who received real stimulation first and those who received sham stimulation first.

Further exploratory analyses were conducted to investigate changes in pre-stimulation pupil size across repeated stimulation trials. These analyses were motivated by the question of whether stimulation-related effects extended to pupil regulation over time. Pre-stimulation pupil size was defined as the mean pupil size during the 500-ms period before stimulation onset. Linear mixed-effects models (*lme4* package) were fitted to account for repeated measurements within participants. The model included condition, group, trial, and their interactions as fixed effects: *pupil ∼ condition × group × trial + (1 | participant).* Random-effects structures were compared using likelihood ratio test and Akaike’s Information Criterion (AIC), with the best-fitting model retained for the final analysis. Estimated marginal trends were calculated using the estimated marginal means (*emmeans* package), with degrees of freedom estimated using Satterthwaite’s approximation.

## 3 Results

### 3.1 Sample characteristics and other outcomes

#### 3.1.1 Sample characteristics

In total, 39 participants (18 healthy and 21 with PD) were recruited for the study. Due to issues during data collection, one participant from the healthy group and one from the PD group was excluded. Two additional participants (one from each group) were excluded after preprocessing because of a high number of trials with substantial data loss. The demographic and clinical characteristics of the final sample are presented in Table 1. No significant group differences were found in age or sex. Stimulation intensity did not differ significantly between groups for real or sham stimulation conditions. Furthermore, paired t-tests revealed no significant differences in stimulation intensity between real and sham conditions in either healthy participants (*t*(15) = -1.90, *p* = 0.1) or patients (*t*(18) = -0.62, *p* = 0.5). Overall, no adverse events were rated as severe and the most common event was a tingling sensation at the ear. A detailed overview of the information obtained using the taVNS-AEQ can be found in the Supplemental Material (Figure S1). Compared with healthy controls, patients with PD showed significantly higher BDI-II scores and lower MoCA and SF-12 mental component scores. No group differences were observed in the IR-REM-BD or SF-12 Physical Component scores. Furthermore, no significant differences were found in reaction times or error rates in the Go/No-Go or Divided Attention tasks. The majority of patients with PD were classified as Hoehn and Yahr stage 1–2, indicating mild disease severity. Consistent with this, MDS-UPDRS scores were generally in the lower range, suggesting that the patient sample was mildly affected. All patients with PD were receiving antiparkinsonian medication and on average took their medication 128 minutes (*SD* = 113.09 minutes) prior to the experiment.

**Table 1.** Demographic, baseline, and clinical characteristics of both groups. Between-group differences were assessed using independent-samples t-tests or Wilcoxon rank-sum tests. Values are presented as mean (*SD*).

| | PD group ($n = 19$) | Healthy controls ($n = 16$) | $p$-value |
| --- | --- | --- | --- |
| Age (years) | 65.3 (7.5) | 66.8 (6.1) | n.s. |
| Sex (f/m) | 7/12 | 8/8 | n.s. |
| MoCA | 25.2 (2.5) | 27.8 (1.8) | **< .01** |
| Stimulation intensity (mA) |  |  |  |
| Real | 2.1 (0.9) | 2.4 (1.1) | n.s. |
| Sham | 2.3 (0.9) | 2.9 (1.0) | n.s. |
| BDI-II | 8.9 (5.62) | 3.4 (3.77) | **< .01** |
| SF-12 |  |  |  |
| Physical component | 44.2 (10.3) | 50.9 (6.58) | n.s. |
| Mental component | 48.9 (9.04) | 56.5 (3.93) | **< .05** |
| IR-REM-BD |  |  |  |
| Patient | 1.5 (3.31) | 0.9 (1.24) | n.s. |
| Other | 1.5 (3.06) | 0.8 (1.09) | n.s. |
| Go/No-Go task |  |  | n.s. |
| Reaction time (ms) | 494 (85.6) | 481 (86.6) | n.s. |
| Error | 0.2 (0.42) | 0.6 (0.89) |  |
| Dual task |  |  |  |
| Reaction time – auditory (ms) | 657 (79.6) | 677 (130) | n.s. |
| Reaction time – visual (ms) | 845 (123) | 857 (120) | n.s. |
| Error | 1.9 (3.59) | 1.7 (2.37) | n.s. |
| MDS-UPDRS |  |  |  |
| Part I | 7.7 (4.50) | - |  |
| Part II | 8.3 (5.41) | - |  |
| Part III | 20.16 (11.84) | - |  |
| Part IV | 2.6 (3.68) | - |  |
| Hoehn & Yahr | 1.9 (0.46) | - |  |
| Time since diagnosis (years) | 4.9 (4.67) | - |  |
| LEDD | 554.9 (264) |  |  |

Note. MoCA, Montreal Cognitive Assessment; BDI-II, Beck's Depression Inventory; SF-12, Short Form 12; IRBD-SSS, International REM Sleep Behavior Disorder Symptoms Severity Scale; MDS-UPDRS, Movement Disorders Society Unified Parkinson's Disease Rating Scale; LEDD, levodopa-equivalent daily dose calculated according to Jost et al., (2023).

### 3.2 Effect of taVNS on pupil size

In the PD group, the cluster-based permutation test revealed one significant cluster in which the pupil dilation was greater in the real than in the sham condition, spanning 2710–3640 ms (*cluster mass* = 252.79, *mean t* = 2.69, *p* = .035). Three additional clusters in the same direction (real > sham) were identified at 1720–1990 ms, 2090–2270 ms, and 3790–3820 ms, but none reached significance (*p* > 0.05). No clusters were identified in the healthy group. The averaged pupil responses for both groups during and after the stimulation are displayed in Figure 2.

**Figure 2.**
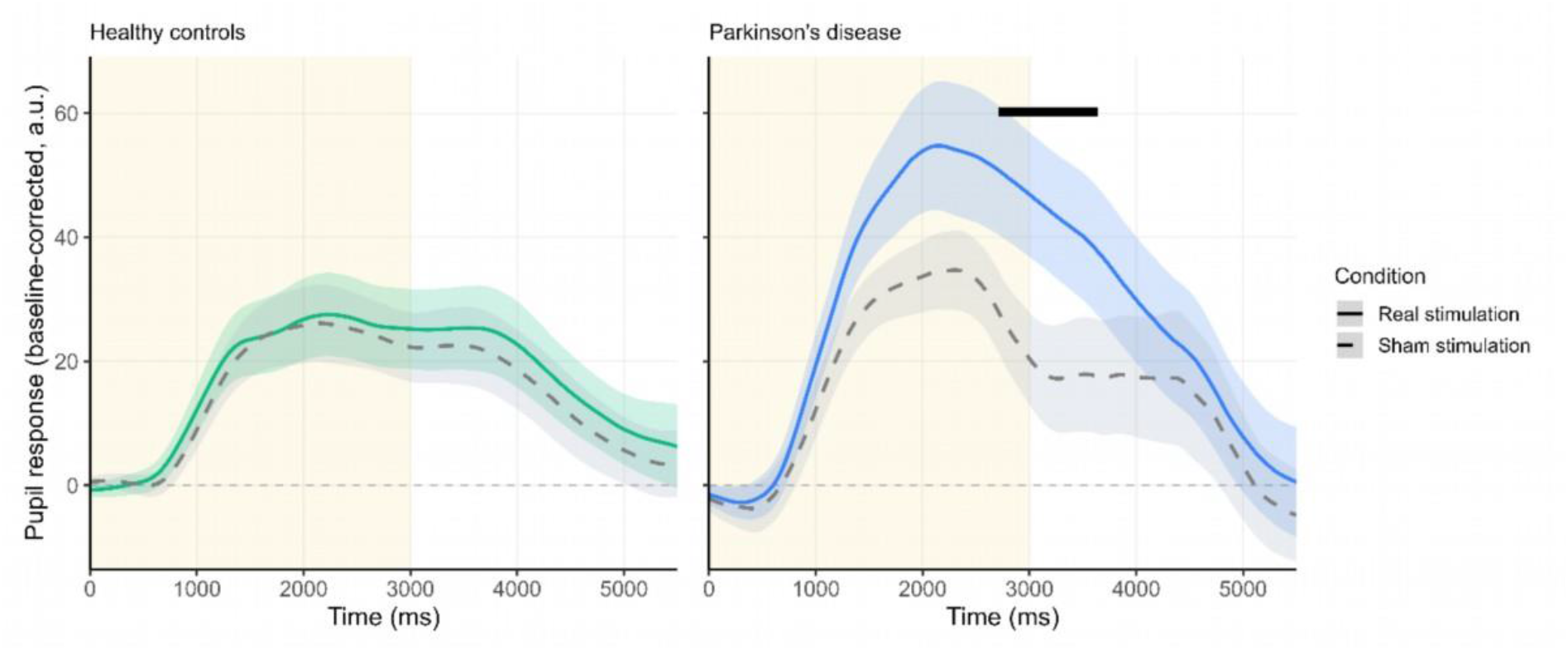
Pupillary responses to real and sham stimulations. Time course of baseline-corrected pupil responses during and after 3 sec of taVNS (indicated by light yellow background). Shaded areas represent ±1 standard error of the mean. Pupil size reported in arbitrary units (a.u.) from the Eyelink eye tracker. The black bar represents the significant cluster found between 2710 and 3640 ms.

Group comparisons of the difference scores (ΔGroup) identified four clusters in which the PD group showed a larger real–sham effect than the healthy control group. Neither cluster reached statistical significance in the cluster-based permutation test. However, a trend toward larger pupil dilation was observed for the PD group compared to healthy controls in a later time window from 2720–3570 ms (*cluster mass* = 179.54, mean *t* = -2.09, *p* = .068). Details of the identified clusters are presented in Tables S1 and S2 in the Supplemental Material. Exploratory analyses on potential condition order effects revealed significant clusters (real_first > sham_first) in the PD group in a window from 570–1440 ms and 1780–3469 ms, Details on the identified clusters can be found in the Supplemental Material (Table S6).

### 3.3 Local-global auditory processing task

In the PD group, the cluster-based permutation test comparing pupil responses to deviant versus non-deviant tones in the local-global auditory processing task revealed one large significant cluster, spanning 490– 2500 ms (*cl.-mass* = 1313.86, *mean t* = 6.50, *p* < .001), in which pupil dilation was greater for deviant than non-deviant stimuli. In the HC group, one significant cluster was found, spanning 460–2500 ms (*cl.-mass* = 1310.48, *mean t* = 6.39, *p* < .001), in which pupil dilation was greater for deviant than non-deviant stimuli. The averaged pupil responses for both groups during the auditory oddball task are displayed in Figure 3.

**Figure 3.**
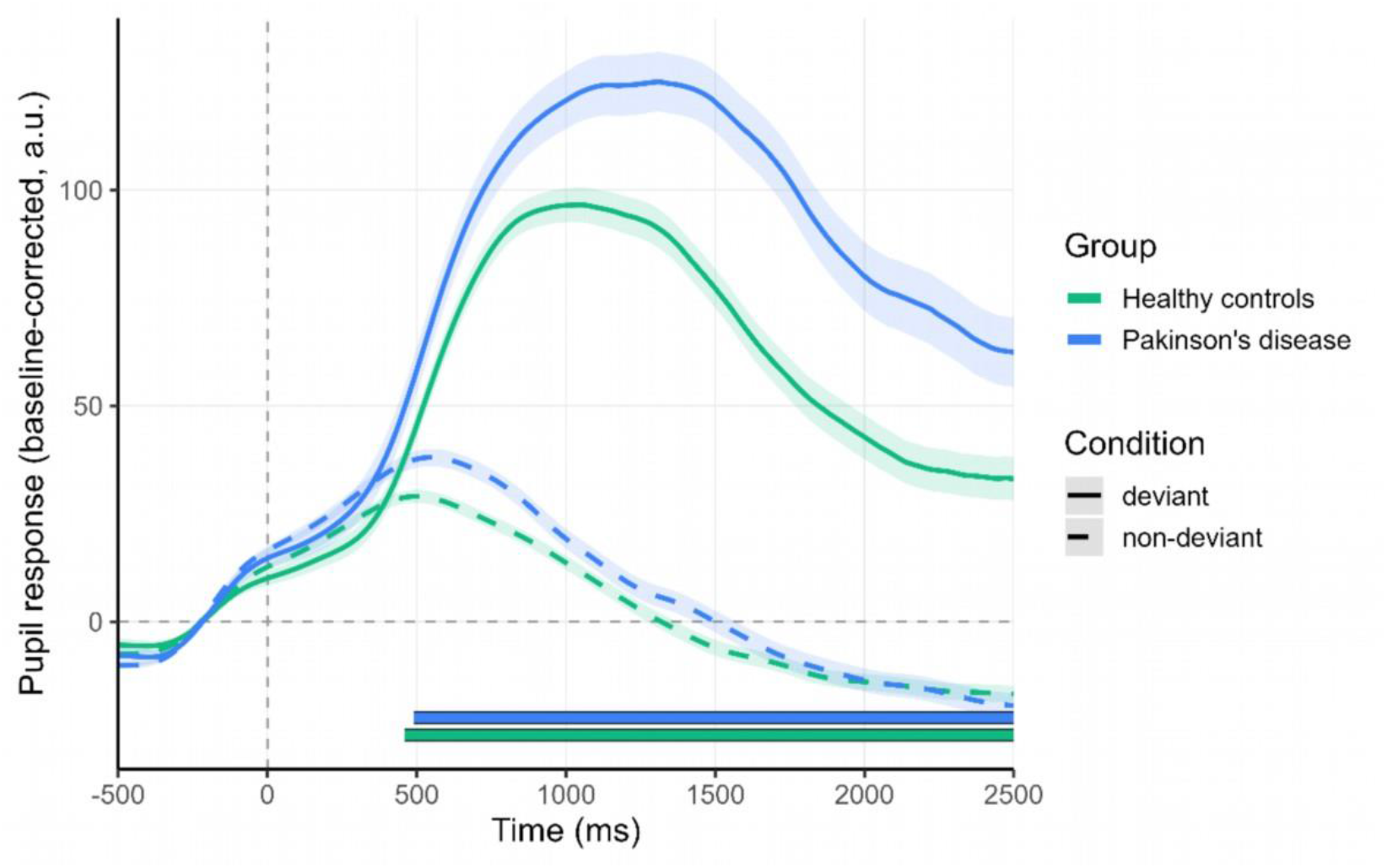
Pupillary responses to deviant and non-deviant tones during the local-global auditory processing task. Time course of baseline-corrected pupil responses following the 5^th^ tone of the tone sequence. Shaded areas represent ±1 standard error of the mean. Pupil size reported in arbitrary units (a.u.) from the Eyelink eye tracker. The blue and green bar represents the significant cluster found in the PD group and the healthy group, respectively.

Group comparisons of the differences scores in the local-global auditory processing task (Δ deviant – non-deviant) revealed a non-significant cluster (1570–1780 ms, *cl.-mass* = 47.41, *mean_t* = -2.15, *p* = .17, PD > Healthy), indicating no statistically significant difference between the PD and healthy groups in the magnitude of deviance-induced pupil response. Details of the identified clusters are presented in the Supplementary Materials in Table S3.

### 3.4 Differences in pre-stimulation pupil size over time

To explore whether pre-stimulation pupil size (i.e., pupil size in the 500 ms window before stimulation onset) changed across trials, a linear mixed-effects model was fitted to examine the effects of condition, group, trial, and their interactions on pre-stimulation pupil size. Adding a random slope for trial significantly improved model fit compared with a random-intercept-only model (*χ²(*2) = 83.38, *p* < .001). Therefore, the model including random intercepts and random slopes for trial was retained as the final model (AIC = 33098; BIC = 33168): *pupil ∼ condition × group × trial + (1 + trial | participant).* In the fixed-effects model, healthy controls and real stimulation were specified as the reference levels for the group and condition, respectively. The trial was z-standardized, such that a value of zero corresponded to the mean trial.

Pre-stimulation pupil size did not differ significantly between stimulation conditions in the control group (*β* = −13.84, *SE* = 9.07, *t*(2563.66) = −1.52, *p* = .128). Participants with PD showed significantly larger pre-stimulation pupil size than healthy controls in the real stimulation condition (*β* = 261.04, *SE* = 86.44, *t(*33.35) = 3.02, *p* = .005). Neither the condition × group interaction, the condition × trial, nor the group × trial interaction reached significance (see Table S5). A significant three-way condition × group × trial interaction (*β* = 41.30, *SE* = 12.51, *t(*2464.62) = 3.30, *p* < .001) was observed, indicating that the effect of trial on pre-stimulation pupil size depended on group and stimulation condition.

Follow-up estimated marginal analyses, with *p*-values adjusted for multiple comparisons using the Holm method, showed a significant decrease in pre-stimulation pupil size across trials in participants with PD during real stimulation (*β* = −42.72, *p* = .001). No significant change across trials was observed in healthy controls during real (*β* = −27.38, *p* = .057) or sham (*β* = −28.28, *p* = .057) stimulation, nor in participants with PD during sham stimulation (*β* = −2.31, *p* = .830). Details are presented in Table S6. Figure 4 displays the averaged pre-stimulation, as well as the estimated marginal means of pre-stimulation pupil size over trials.

**Figure 4.**
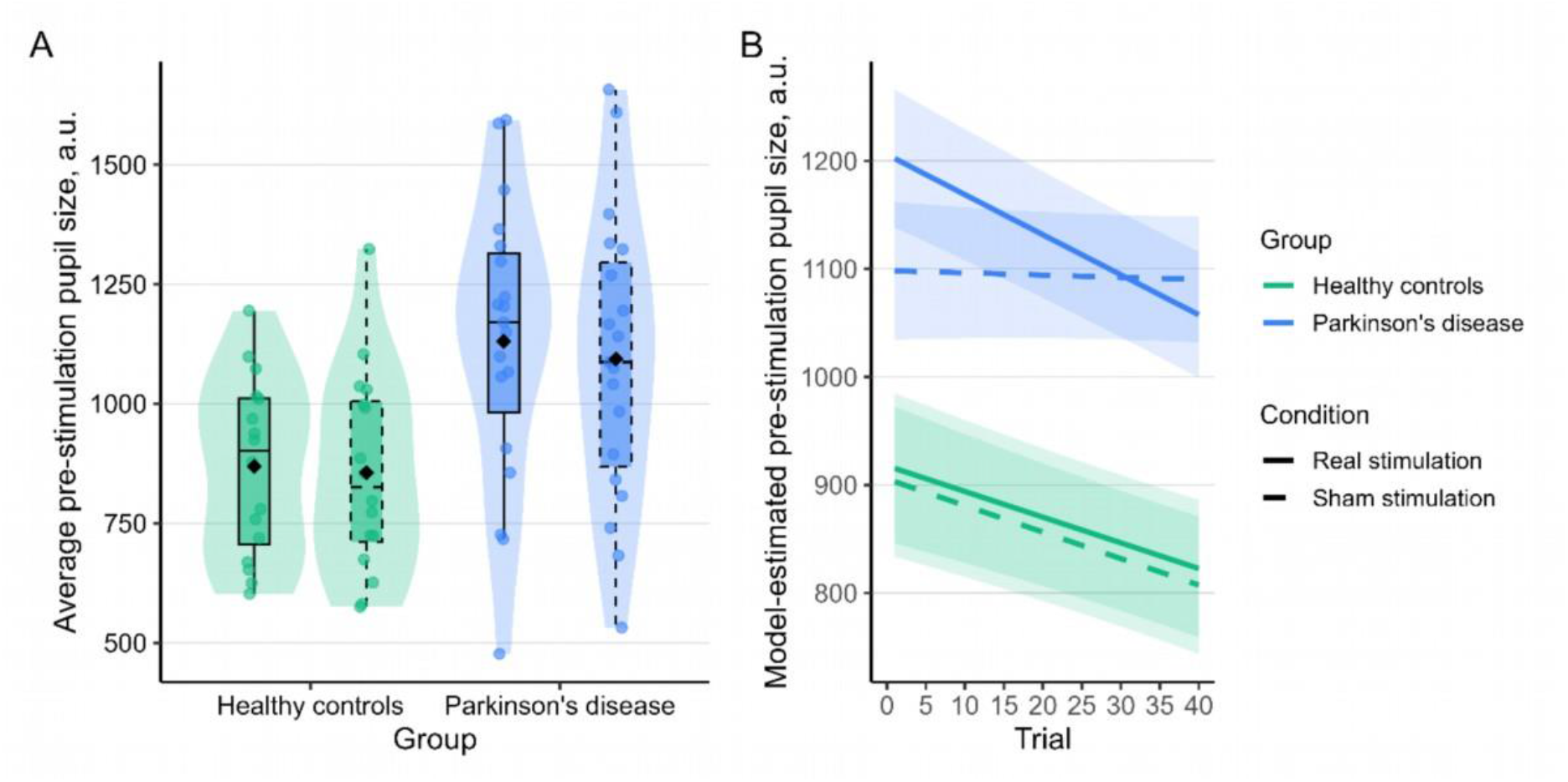
Pre-stimulation pupil size across groups and stimulation conditions. (A) Average pre-stimulation pupil size by group and stimulation condition. The points represent individual participants; black diamonds indicate group means. (B) Estimated marginal means from the linear mixed-effects model showing pre-stimulation pupil size across trials for both conditions and groups. Shaded ribbons represent ±1 standard error. Pupil size reported in arbitrary units (a.u.) from the Eyelink eye tracker.

## 4 Discussion

This study investigated whether taVNS modulates pupil responses in PD. The pupil response was used as a non-invasive indirect marker of neuromodulatory or arousal-related activity closely related to theLC-NE activity. Compared with sham stimulation, real taVNS led to significantly greater pupil dilation in the PD group, with a significant cluster emerging between 2710 and 3640 ms after the stimulation onset. In contrast, no significant difference between real and sham stimulation was found in the healthy control group. A direct comparison of the taVNS-induced pupil response between groups did not reveal a significant group difference. Exploratory analyses of potential order effects revealed two significant clusters in the PD group, with greater responses when real stimulation was administered first compared with when sham was administered first. Furthermore, exploratory linear-mixed effects models indicated that changes in pre-stimulation pupil size across trials depended on stimulation condition and group. Finally, the local-global auditory processing task effectively elicited larger pupil responses to deviant tones compared to non-deviant tones in both groups. This finding indicates that both groups were capable of generating arousal-related responses, and the associated pupil responsiveness may reflect locus coeruleus-norepinephrine (LC-NE) activity.

### 4.1 taVNS modulates pupil responses in PD, not in healthy controls

The findings indicate that brief (3 s) taVNS can lead to measurable changes in pupil size in PD, suggesting that vagal stimulation can elicit rapid autonomic responses in this group. The present study was designed to investigate the immediate physiological response to a brief stimulation to increase the understanding of how taVNS affects the pupil response as a proxy for neuromodulatory or arousal-related activity related to the LC-NE system in PD, where degeneration of the LC and alterations in noradrenergic signalling have been implicated in disease pathology (Braak et al., 2004; Paredes-Rodriguez et al., 2020). Previous studies have reported improvements in motor and non-motor symptoms following taVNS in PD (Proietti et al., 2025), although these studies have generally used longer stimulation protocols as their findings primarily address the potential clinical effects of the stimulation. The mechanisms underlying the effects of taVNS remain largely unexplored in existing studies. Our study has advanced the understanding of these stimulation-associated effects by demonstrating that taVNS influences the brainstem, likely through the LC-NE system, as evidenced by pupil dilation observed in this study. A stimulation duration of 3 seconds was employed, shorter than that used in clinical studies, as brief stimulation is adequate for detecting pupil effects (Skora et al., 2024)

The absence of a significant taVNS-related pupil response in healthy controls may be explained by age-related changes in pupillary and autonomic functions. Age-related miosis and alterations in LC structure and function can potentially affect the magnitude of pupil responses to vagal stimulation (Alrosan et al., 2024; Fotiou et al., 2007; Parashar, 2016, 2016; Riley et al., 2025; Sloane et al., 1988). However, these changes are also relevant to the PD group, as participants with PD were in the same age range. Thus, age-related factors alone are unlikely to explain the observed pattern, disease- or arousal-related alterations in neural and autonomic systems involved in pupil regulation may also contribute. Importantly, both healthy controls and participants with PD showed clear pupil responses to deviant auditory stimuli in the local-global auditory processing task, confirming preserved stimulus-evoked neuromodulatory pupil responsiveness in both groups. This suggests that the absence of a significant taVNS-related response in healthy controls was not simply due to a generally reduced NE driven pupil responsivity. Age- and disease-related changes in LC structure and function may contribute to the observed pattern, as previous studies have reported changes in LC integrity and evidence for potential compensatory mechanisms (Ludwig, Yi, et al., 2024; Paredes-Rodriguez et al., 2020; Riley et al., 2025). However, differences in the initial arousal or neuromodulatory states may also be relevant. Recent evidence suggests that the effects of LC-NE activity on functional brain dynamics are not necessarily linear, but can depend on baseline arousal, with an inverted U-shaped relationship between arousal and functional connectivity (Tong et al., 2025). Thus, the observed taVNS-related pupil response in PD, in contrast to the absence of a significant response in healthy controls, could reflect differences in participants’ initial arousal or neuromodulatory state. In this context, both disease-related changes and state-dependent neuromodulatory effects may have contributed to the observed pattern. Further work directly assessing baseline arousal and LC-NE function is necessary to disentangle these potential contributions.

### 4.2 Effects on pre-stimulation pupil size in PD

Overall, patients with PD showed larger pre-stimulation pupil sizes than healthy controls, which may partly reflect effects of dopaminergic medication (Bartošová et al., 2025), but could potentially also indicate alterations in arousal or neuromodulatory systems, including LC-NE-related activity. Differences in pre-stimulation pupil size over time may reflect the effect of taVNS in PD beyond the immediate taVNS-induced pupil response found in this study. Baseline pupil size is associated with tonic LC activity and arousal (Joshi et al., 2016; Joshi & Gold, 2020). Across trials, pre-stimulation pupil size showed a significant decrease only in patients with PD during real taVNS, whereas no significant trial-related changes were observed in the other conditions. Notably, the estimated slopes were negative in both stimulation conditions in healthy controls and during real taVNS in patients with PD, while the slope was close to zero during sham stimulation in patients with PD. This pattern suggests that the temporal change in pre-stimulation pupil size may differ between stimulation conditions in PD. The present analyses do not allow these changes to be specifically attributed to LC-NE activity and may instead reflect alterations in arousal-related or autonomic processes more broadly. Therefore, future studies should examine changes in pre-stimulation pupil size across repeated taVNS trials more systematically, ideally alongside additional measures of autonomic and LC-related activity, to determine whether such changes reflect sustained effects of taVNS on arousal or LC function in PD.

### 4.3 Different pathways underlying LC-related pupil responses

The different pupillary responses observed across paradigms may reflect differences in the physiological pathways engaged by auditory stimuli and taVNS. While both paradigms are associated with LC-NE-related pupil responses, they engage the system through distinct afferent pathways. Auditory deviants naturally engage the LC-NE system, whereas taVNS may activate it through external electrical stimulation. The less pronounced and slightly delayed pupil response observed during taVNS compared with the local-global auditory processing response (see Figures 2 and 3) may be indicative of these differences. The preserved auditory-evoked pupil response in both groups, contrasted with the taVNS-related response observed only in PD, may therefore suggest that disease-related alterations or compensatory mechanisms specifically within the vagal-brainstem pathway may contribute to the observed pattern.

### 4.4 Limitations and future directions

The interpretation of the present findings is supported by previous research linking pupil dynamics to LC activity (Gilzenrat et al., 2010; Murphy et al., 2014; Reimer et al., 2016). However, pupillometry remains an indirect measure of LC activity and is influenced by multiple neural and physiological factors (Joshi et al., 2016; Joshi & Gold, 2020; Mazancieux et al., 2023; Szabadi, 2018). Moreover, taVNS does not exclusively target the LC, but may also modulate other neurotransmitter systems (Proietti et al., 2025). Combining pupillometry with additional measures, such as functional or neuromelanin-sensitive MRI, could provide more detailed evidence of LC integrity and activity and help clarify the mechanisms underlying taVNS-induced pupil responses in PD.

Another potential limitation of the present study is the relatively short washout period between stimulation conditions. The subsequent stimulation condition was administered 10 minutes after the preceding one. Prior stimulation may affect the pupillary response, future studies should therefore implement longer washout periods to minimize such effects and further investigate the duration required to adequately prevent them.

The relatively small sample of predominantly mildly affected individuals limit the generalizability of the findings across disease stages. Further investigation of whether taVNS responsivity varies with disease stage and subtype, and whether individual differences in vagal integrity (Laucius et al., 2025) or sensitivity to stimulation (Ludwig, Pereira, et al., 2024) contribute to its variability is needed. Longitudinal studies could additionally determine whether repeated taVNS induces sustained changes in physiological measures related the LC-NE system and whether these changes are associated with improvements in motor and, particularly, non-motor symptoms in PD.

The few and mild adverse events observed in the present study are consistent with the tolerability and feasibility of taVNS in PD (Lench et al., 2023), supporting its potential as a non-invasive neuromodulatory approach. Overall, the present findings provide initial evidence that brief taVNS can modulate pupil responses in PD, consistent with the possibility that LC-NE-related processes are involved. Further research is needed to clarify the underlying mechanisms and determine whether these neuromodulatory effects can have therapeutic relevance.

### Authors’ Roles

(1) Research Project: A. Conception, B. Organization, C. Execution; (2) Data Analysis: A. Design, B. Execution, C. Review and Critique; (3) Manuscript Preparation: A. Writing of the First Draft, B. Review and Critique.

L.M.S.: 1A, 1B, 1C, 2A, 2B, 3A

J.S.: 1B, 2C, 3B

C.M.T.: 1A, 2C, 3B

K.J.: 1B, 2C, 3B

K.W.: 1A, 2B 2C, 3A

## Acknowledgment

We would like to thank all study participants for taking part in the study. We also thank Svenja Schwichtenberg for allowing us to use the local-global auditory processing task and Angelina Maikranz and Sünje Andermann for their help with data collection and data management.

## Data availability statement

The data presented in this study are available on request from the corresponding author.

