## Supplemental Material for "Transcutaneous auricular vagus nerve stimulation demonstrates effects on pupil responses in Parkinson’s disease"

### Supplement 1 - Adverse events of the stimulation

The taVNS-AEQ indicated that the most frequent adverse event was a tingling sensation at the ear, experienced by over 80% of participants in both conditions, followed by tiredness, which was reported by over 40% of participants in the real condition and over 50% in the sham condition. In the sham condition, participants reported between 0 and 5 adverse events, with most participants experiencing three events (*n* = 12), followed by one and two events (*n* = 8). Under real stimulation condition, the number of reported adverse events ranged from 0 to 6. The most common was one reported event (*n* = 12), followed by three and two events (*n* = 8 and *n* = 6, respectively). No adverse events were rated as severe or highly distracting. No participant discontinued the study due to stimulation-related effects, and all participants indicated their willingness to participate in future studies involving stimulation in both real and sham conditions. Overall, 50% of participants correctly identified their stimulation condition, indicating a chance level. An overview of adverse events reported in this study is provided in Figure S1.

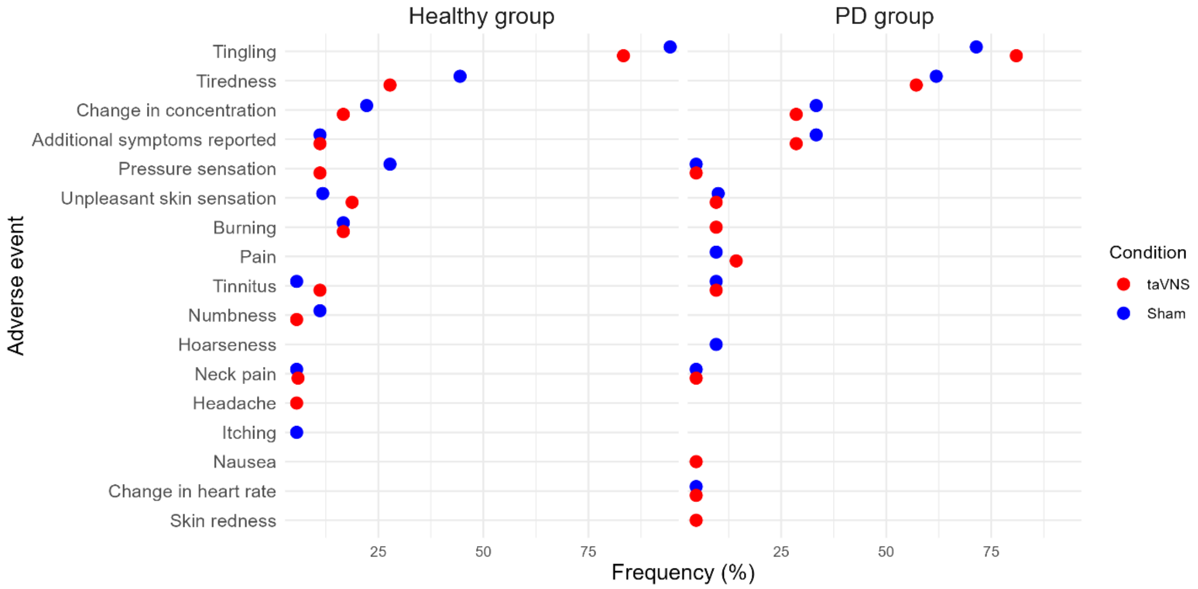


**Figure S1.** Frequency of adverse events during and after stimulation under both stimulation conditions and for both groups.

### Supplement 2 – Details on the cluster permutation analyses

**Table S1.** Identified clusters for taVNS in the PD group.

| **Cluster** | **Time (ms)** | **Mass** | **Mean *t*** | ***p*-value** | **Direction** |
|---|---|---|---|---|---|
| 1 | 1720–1990 | 64.00 | 2.29 | .229 | Real > Sham |
| 2 | 2090–2270 | 42.38 | 2.23 | .297 | Real > Sham |
| 3 | 2710–3640 | 252.79 | 2.69 | **.034** | Real > Sham |
| 4 | 3790–3820 | 8.47 | 2.12 | .479 | Real > Sham |

*Note*. Cluster-based permutation test (two-sided, sign-flipping, 10,000 permutations) comparing real vs. sham pupil responses in the PD group. Cluster 3 survived correction for multiple comparisons ($p < .05$).

**Table S2.** Identified clusters for taVNS, group differences.

| **Cluster** | **Time Window (ms)** | **Mass** | **Mean *t*** | ***p*-value** | **Direction** |
|---|---|---|---|---|---|
| 1 | 1680–2010 | 70.75 | -2.08 | .198 | PD > Healthy |
| 2 | 2420–2470 | 10.54 | -1.76 | .467 | PD > Healthy |
| 3 | 2510–2520 | 3.45 | -1.73 | 0.522 | PD > Healthy |
| 4 | 2720–3570 | 179.54 | -2.09 | 0.068 | PD > Healthy |

*Note*. Two-sample cluster-based permutation test (one-sided, label-shuffling, 10,000 permutations) comparing healthy vs. PD difference scores (Δstim = real − sham). No clusters survived correction for multiple comparisons.

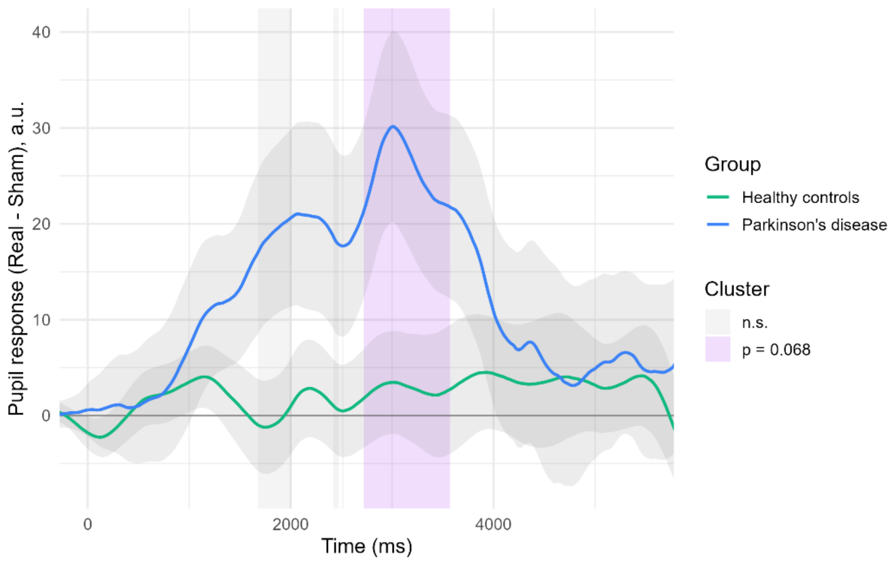


**Figure S2.** Mean pupil response difference (Real – Sham condition) over time for both groups with shaded bands representing ±1 SEM. The purple-shaded time window indicates the cluster showing a trend-level group difference (PD > Healthy). Pupil size reported in arbitrary units (a.u.) from the Eyelink eye tracker.

**Table S3.** Identified clusters of pupil dynamics for the local-global auditory processing task in the healthy control and PD group.

| **Group** | **Cluster** | **Time (ms)** | **Mass** | **Mean *t*** | ***p*-value** | **Direction** |
|---|---|---|---|---|---|---|
| HC | 1 | 230–330 | 25.71 | −2.34 | .195 | Non-deviant > Deviant |
| | 2 | 450–2500 | 1186.61 | 5.76 | **< .001** | Deviant > Non-deviant |
| PD | 1 | 480–2500 | 1339.34 | 6.60 | **< .001** | Deviant > Non-deviant |

*Note*. Cluster-based permutation tests (two-sided, sign-flipping, 10,000 permutations) comparing pupil responses to deviant vs. non-deviant stimuli, conducted separately for the HC and PD groups. One significant cluster per group was found.

### Supplement 3 - Exploratory analyses of potential carry-over effects

Potential carry-over effects were analysed exploratively to observe whether the order of stimulation condition received (real or sham stimulation first) affected the difference between real and sham stimulation conditions.

Cluster-based permutation analyses for the difference between Δreal_first (= real-sham) and Δsham_first (= real-sham) were conducted separately for both groups.

**Table S4.** Identified clusters for the order effect on the difference between Δreal_first and Δsham_first for the PD group.

| Cluster | Time (ms) | Mass | Mean *t* | *p*-value | Direction |
|---|---|---|---|---|---|
| **1** | **570–1440** | **258.26** | **2.93** | **.036** | **RealFirst > ShamFirst** |
| **2** | **1780–3460** | **510.36** | **3.02** | **.010** | **RealFirst > ShamFirst** |
| 3 | 3500–3640 | 33.21 | 2.21 | .370 | RealFirst > ShamFirst |
| 4 | 4250–4530 | 64.43 | 2.22 | .252 | RealFirst > ShamFirst |
| 5 | 4550–4550 | 2.11 | 2.11 | .574 | RealFirst > ShamFirst |

In patients with PD, the difference between real and sham conditions was larger in individuals who received real stimulation first than in those who received sham first. This was evident for an early window (Cluster 1) and most importantly for a window (Cluster 2) falling into the time-span of the cluster observed in the main analysis.

In healthy controls, no clusters were identified in the comparison of the difference (real-sham) between individuals receiving real stimulation or sham stimulation first.

### Supplement 4 – Details on the exploratory analysis of pre-stimulation pupil size

**Table S5.** Linear mixed-effects model predicting pre-stimulation pupil size.

| **Fixed effect** | ***β*** | ***SE*** | ***df*** | ***t*** | ***p*** |
|---|---|---|---|---|---|
| Intercept | 869.34 | 63.68 | 33.34 | 13.65 | **<.001** |
| Condition (Real vs. Sham) | −13.84 | 9.09 | 2463.66 | −1.52 | .128 |
| Group (PD vs. Healthy) | 261.04 | 86.44 | 33.35 | 3.02 | **.005** |
| Trial | −27.38 | 11.64 | 46.61 | −2.35 | **.023** |
| Condition x Group | −22.28 | 12.47 | 2463.65 | −1.79 | .074 |
| Condition x Trial | −0.89 | 9.22 | 2463.88 | −0.10 | .923 |
| Group x Trial | −15.33 | 15.78 | 46.49 | −0.97 | .336 |
| Condition x Group x Trial | 41.30 | 12.51 | 2464.62 | 3.30 | **<.001** |

*Note.* Healthy controls and real stimulation were specified as the reference levels. Trial was z-standardized. The model included random intercepts and random slopes for trial by participant: baseline ~ condition × group × trial + (1 + trial | participant). Degrees of freedom and tests were based on the Satterthwaite approximation.

**Table S6.** Follow up estimated marginal trends of trial by group and stimulation condition.

| **Group** | **Condition** | **Trial slope (*β*)** | ***SE*** | ***df*** | ***t*** | ***p*-adj.** | **95% CI** |
|---|---|---|---|---|---|---|---|
| Healthy | Real | -27.38 | 11.64 | 46.60 | −2.35 | .057 | [−50.80, −3.97] |
| | Sham | −28.28 | 11.64 | 46.46 | −2.43 | .057 | [−51.70, −4.88] |
| PD | Real | −42.72 | 10.67 | 46.27 | −4.01 | **.001** | [−64.20, −21.25] |
| | Sham | −2.31 | 10.70 | 46.70 | −0.22 | .830 | [−23.80, 19.19] |

*Note.* Trial slopes represent changes in pre-stimulation pupil size per 1 SD increase in trial number. P-values were adjusted for four follow-up tests using the Holm method. Confidence intervals shown are 95% CIs.
